# Identifying subtypes of heart failure with machine learning: external, prognostic and genetic validation in three electronic health record sources with 320,863 individuals

**DOI:** 10.1101/2022.06.27.22276961

**Authors:** Amitava Banerjee, Suliang Chen, Muhammad Dashtban, Laura Pasea, Johan H Thygesen, Ghazaleh Fatemifar, Benoit Tyl, Tomasz Dyszynski, Folkert W. Asselbergs, Lars H. Lund, Tom Lumbers, Spiros Denaxas, Harry Hemingway

**Affiliations:** Institute of Health Informatics, University College London, London, UK; Health Data Research UK, University College London, London, UK; Barts Health NHS Trust, London, UK; University College London Hospitals NHS Trust, London, UK; Bayer HealthCare SAS, Medical Affairs, Pharmaceuticals, BP 103 10 Place de Belgique, F-92254 La Garenne Colombes Cedex, France; Bayer AG, Medical Affairs & Pharmacovigilance, Pharmaceuticals TG Cardio, Thrombosis & Hemophilia Building M084, 112 13353 Berlin, Germany; Department of Cardiology, Division Heart & Lungs, University Medical Center Utrecht, Utrecht University, Utrecht, the Netherlands; Institute of Cardiovascular Science, Faculty of Population Health Sciences, University College London, London, United Kingdom; Division of Cardiology, Department of Medicine, Karolinska Institutet, Stockholm, Sweden; Heart and Vascular Theme, Karolinska University Hospital, Stockholm, Sweden; University College London Hospitals National Institute for Health Research (NIHR) Biomedical Research Centre

## Abstract

**Background:** Reliable identification of heart failure (HF) subtypes might allow targeted management. Machine learning (ML) has been used to explore HF subtypes, but neither across large, independent, population-based datasets, nor across the full spectrum of causes and presentations, nor with clinical and non-clinical validation by different ML methods. Using our published framework, we identified and validated HF subtypes to address these gaps.

**Methods:** We analysed individuals ≥30 years with incident HF from two population-based electronic health records resources (1998-2018; Clinical Practice Research Datalink, CPRD: n=188,799 HF cases; The Health Improvement Network, THIN: n=124,263 HF cases). Pre-and post-HF factors (n=645) included demography, history, examination, blood laboratory values and medications. We identified subtypes using four unsupervised ML methods (K-means, hierarchical, K-Medoids and mixture model clustering) with 87 (from 645) factors in each dataset. We evaluated subtypes for: (i) *external validity* (across independent datasets); *(ii) prognostic validity* (predictive accuracy for 1-year mortality); and (iii) uniquely, *genetic validity* (in UK Biobank; n=9573 cases): association with polygenic risk score (PRS) for 11 HF related traits, and direct association with 12 reported HF single nucleotide polymorphisms (SNPs).

**Findings:** After identifying five clusters, we labelled HF subtypes: 1.Early-onset, 2.Late-onset, 3.AF-related, 4.Metabolic, and 5.Cardiometabolic. *External validity:* Subtypes were similar across datasets (c-statistic: 0.94, 0.80, 0.79, 0.83, 0.92 for the THIN model in CPRD and 0.79, 0.92, 0.90, 0.89, 0.92 for the CPRD model in THIN for subtypes 1-5, respectively). *Prognostic validity:* One-year all-cause mortality, risk of non-fatal cardiovascular diseases and all-cause hospitalisation (before and after HF diagnosis) differed across subtypes in CPRD and THIN data. *Genetic validity:* The AF-related subtype showed associations with PRS for related traits. Late-onset and Cardiometabolic subtypes were most comparable and strongly associated with PRS for Hypertension, Myocardial Infarction and Obesity (p-value < 9.09 × 10^−4^). We developed a prototype for clinical use, which could enable evaluation of effectiveness and cost-effectiveness.

**Interpretation:** Across four methods and three datasets, and including genetic data, in the largest HF study to-date, ML algorithms identified five subtypes in individuals with incident HF. These subtypes may inform aetiologic research, clinical risk prediction and the design of HF trials.

**Funding:** European Union Innovative Medicines Initiative.

**Research in context:** 

**Evidence before this study:** In a systematic review until December 2019, we showed that studies of machine learning in subtyping and risk prediction in cardiovascular diseases are limited by small population size, relatively few factors and poor generalisability of findings due to lack of external validation. We further searched PubMed, medRxiv, bioRxiv, arXiv, for relevant peer-reviewed articles and preprints, focusing on machine learning studies in heart failure. Studies remain focused on single diseases, limited risk factors, often single method of machine learning, rarely use subtyping and risk prediction together, and have not been externally validated across datasets. For heart failure, all subtype discovery studies have identified subtypes based on clustering, but so far with no application to clinical practice.

**Added value of this study:** Across two independent, population-based datasets, we used four machine learning methods for subtyping and risk prediction with 89 aetiologic factors as well as 556 further factors for heart failure. We identified and validated five subtypes in incident heart failure, which differentially predicted outcomes. In addition, we externally validated clinical cluster differences by exploring corresponding genetic differences in a large-scale genetic cohort. Our methods and results highlight potential value of electronic health records and machine learning in understanding disease subtypes. Moreover, our approach to external, prognostic, and genetic validity provides a framework for validation of machine learning approaches for disease subtype discovery.

**Implications of all the available evidence:** Our analyses support coordinated use of large-scale, linked electronic health records to identify and validate disease subtypes with relevance for clinical risk prediction, patient selection for trials, and future genetic research.

## Introduction

### Importance

Heart failure (HF) is a heterogeneous syndrome reflecting multiple underlying causes (European Society of Cardiology, ESC: 13 categories and 89 individual causes)(1). Disease subtypes may be relevant, where single aetiologic factors in isolation (e.g. diabetes, myocardial infarction (MI)) have not necessarily improved characterisation of HF diagnosis(2) or prognosis, discovery of new treatments, trial design or clinical decision-making(3), despite causal associations(4, 5). Current subtype classifications, including by aetiology (e.g. ischaemic vs non-ischaemic), pathophysiology (e.g. primary myocardial disease vs secondary neurohormonal activation), anatomy (e.g. left-vs right-sided(6)), haemodynamics (hypoperfusion vs congestion), presentation (e.g. acute vs chronic(6, 7)), setting (e.g. outpatient vs inpatient), left ventricular ejection fraction, LVEF (e.g. reduced vs mid-range or mildly reduced vs preserved(1)), symptoms (e.g. New York Heart Association class 1-4(8) or American Heart Association HF stage A-D(9)), comorbidities (e.g. end-stage renal disease(10)) or biomarkers (e.g. NT-proBNP (N-terminus-pro-brain natriuretic peptide) (9)), have not led to “precision medicine,” “personalised care,” or targeted therapies. Incomplete knowledge of subtypes across the whole range of aetiologic factors and population has also limited primary prevention and screening guidelines for HF(10, 11).

In clinical practice and research, subtypes are commonly classified by LVEF for diagnosis and prognosis, but in a recent machine learning (ML) study, EF did not predict survival in a large HF registry(12). ML is rarely used to identify subtypes in large, nationally representative datasets linked across healthcare settings (i.e. primary and secondary care), where “agnostic”, unsupervised subtype discovery across risk factors may inform HF treatment and prevention. Moreover, studies to improve HF subtype classification and risk prediction have neither compared different ML methods in one study nor validated ML-based subtypes in a separate population, with limited studies of risk prediction. Our seven-stage framework (clinical relevance, patients, algorithm, internal validation, external validation, clinical utility, effectiveness) for practical ML implementation may yield more clinically relevant results (13).

Therefore, in a population of 322,635 individuals with incident HF and 645 factors across three population-based datasets, we used four unsupervised ML methods to:

(i) Generate subtypes with clinical relevance throughout the course of HF, and low risk of bias for patient selection and algorithms (<u>“</u>*<u>Development”</u>*).
(ii) Demonstrate validity: internal (across methods), external (across datasets), prognostic (predictive accuracy for 1-year all-cause mortality) and genetic (using known single nucleotide polymorphisms, SNPs, associated with HF) *<u>(“Validation”)</u>*.
(iii) Propose methods to improve impact (clinical utility and effectiveness) *<u>(“Impact”)</u>*.

## Methods

We used our published framework for ML implementation to inform our methods (13).

### (1) To generate subtypes (Development)

#### Clinical relevance

By aiming to improve diagnostic and prognostic prediction of HF, our research concerned “patient benefit”. We used two independent population-based primary care electronic health records (EHRs) with validity for HF and cardiovascular diseases (CVD) research (“target condition applicability”: whether the disease defined in data matches research questions). Primary care EHR (The Health Improvement Network, THIN(14) and Clinical Practice Research Datalink, CPRD-GOLD), were linked by CPRD and NHS Digital (using unique national healthcare identifiers), with hospital admissions (Hospital Episodes Statistics, HES), and death registry (Office for National Statistics, ONS)(15). Both datasets are representative of the UK population, with prospective recording and follow-up (“data suitability”). For genetic validation, we used UK Biobank data (UKB)(16), comprising initial release of genotyping for a random sample of 150,000 of 502,641 participants, aged 40-69 years (recruited 2006-2010), linked to primary care (∼50%) and secondary care (100%).

#### Patients

We ensured “patient applicability” (to study aims), minimising patient selection bias, including individuals ≥30 years with incident HF (1 January 1998-1 January 2018) and ≥1 year of follow-up in CPRD and THIN. Given overlap between THIN and CPRD practices, we avoided double counting individuals using validated methods(17, 18)(**Web Figure 1**). We defined incident HF as first record of fatal or non-fatal, hospitalised or non-hospitalised HF in primary (Read coding) or secondary care (International Statistical Classification of Diseases-10th version, ICD-10) based on HDR-UK CALIBER phenotypes (**Web Figure 2 and 3**)(19).

#### Algorithm

We included 645 factors: (i)demography(e.g. age; n=16); (ii)aetiology based on ESC classification(n=258)(1, 11); (iii)comorbidities (e.g. depression; n=114); (iv)symptoms (e.g. dyspnoea; n=39); (v)medication use and persistence (by 90-day prescription gap over 1 year)(HF and non-HF; n=84); (vi)examination (e.g. blood pressure; n=11); (vii)investigations (e.g. kidney function; n=24); and (viii)non-CVD factors, based on a prior ML study(n=99)(20)(**Web Table 1)**. Existing phenotypes were used if possible and new phenotypes (n=23) were developed using a standardised, rule-based approach(15). Factors were classified as “before” (in the 5 years prior to or at time of HF diagnosis, e.g. prior ACE-inhibitor), “after” (during follow-up, e.g. ACE-inhibitor post-HF diagnosis) or “ever” (before or after). Like prior studies(19), use of pre-and post-HF factors maximised available data use in a given individual, and clinical and research utility over the course of HF (i.e. not just baseline).

Factors with >30% missing data were excluded from clustering analyses(n=10). In remaining continuous and categorical factors, we imputed missing data by Principal Component Analysis (PCA: timed scores regression) and Multiple Correspondence Analysis (MCA) respectively. For dimensionality reduction, we used Random Forest supervised classification for 1-year mortality, ranking variable importance by prediction accuracy and Gini coefficient(21)(**Figure 1, Web Figure 4**). Risk of algorithmic bias was minimised by comparing four ML methods: K-means (partitioning, non-parametric), Hierarchical (agglomerative), K-medoids (partitioning and dissimilarity matrix, and non-parametric) and Mixture modelling (parametric).

**Figure 1.**
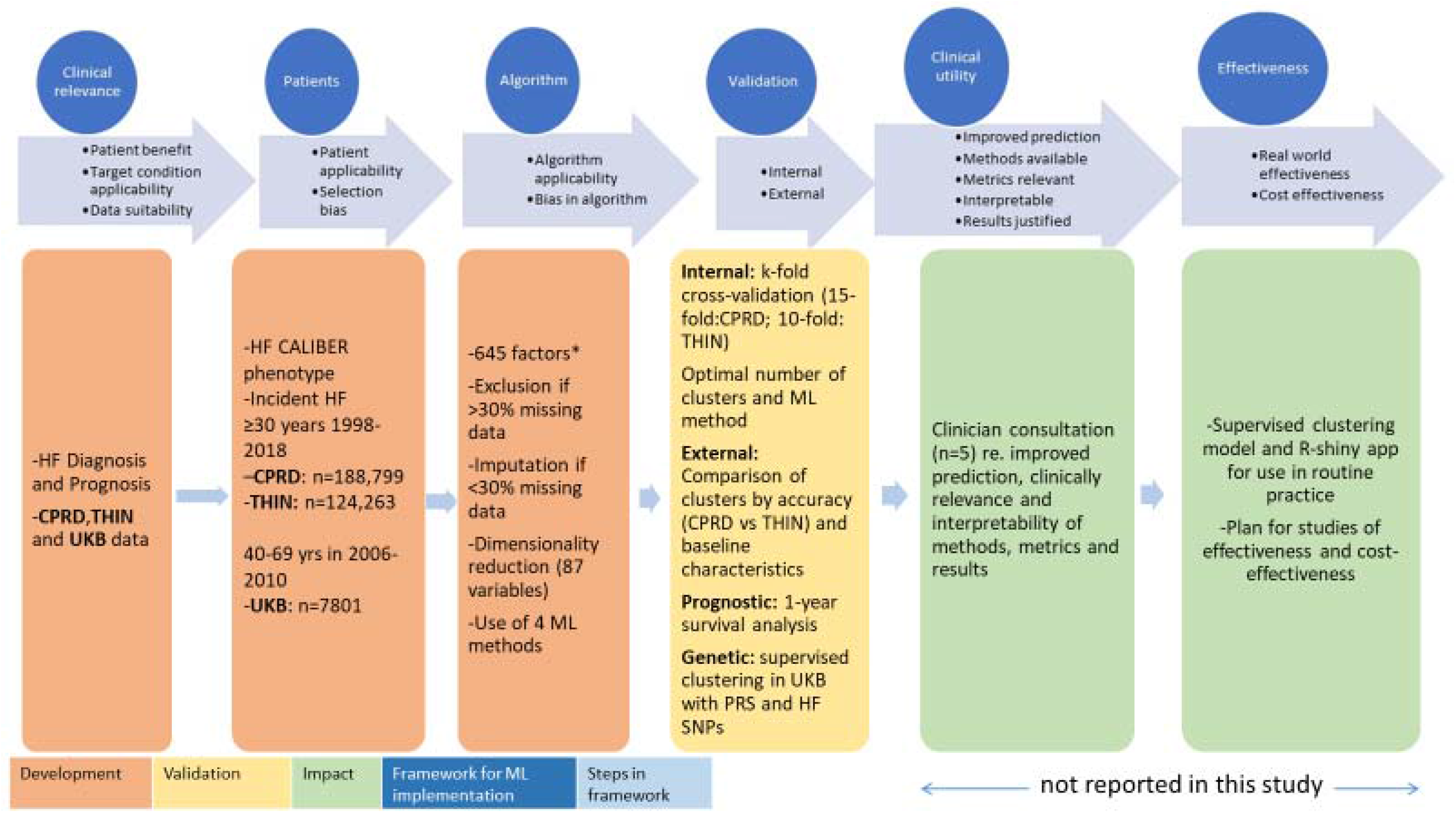
Study design for development, validation, and evaluation of impact of machine learning-led subtyping in incident heart failure. CPRD: Clinical Practice Research Datalink; THIN: The Health Improvement Network. 645 factors included: (i)demography(e.g. age; n=16); (ii)aetiology based on ESC classification(n=258)(1, 11); (iii)comorbidities (e.g. depression; n=114); (iv)symptoms (e.g. dyspnoea; n=39); (v)medication use and persistence (by 90-day prescription gap over 1 year)(HF and non-HF; n=84); (vi)examination (e.g. blood pressure; n=11); (vii)investigations (e.g. kidney function; n=24); and (viii)non-CVD factors, based on a prior ML study(n=99)

### To demonstrate validity (Validation)

#### Internal (within dataset and across methods)

After training and validation (CPRD: 15-fold; THIN: 10-fold), one fold was selected to represent the whole population in each dataset. PCA reduced noise, variance and dimensionality. We determined optimal number of clusters (silhouette width, prediction strength(22)) and ML method (similarity indices and matrices, and cluster stability: Rand, Jaccard means and Fowlkes and Mallows’ indices(23))(**Web Table 2**).

#### External (across datasets)

Clusters were compared by accuracy between datasets (e.g. CPRD clusters predicting clusters in THIN), and baseline continuous (means: analysis of variance, ANOVA) and categorical (proportions: Pearson’s χ2 test) factors(**Web Table 3**).

#### Prognostic (predictive accuracy for 1-year all-cause mortality)

We analysed prevalence and incidence for risk factors diseases and drugs in each CPRD and THIN cluster before and after incident HF, comparing Kaplan-Meier 1-year survival (log-rank for differences; p<0.01). *Genetic (polygenic risk scores, PRS, and single nucleotide polymorphisms, SNP):* Using identified cluster labels, we built a supervised learning model to predict clusters in those with HF. We assessed cross-cluster genetic differences via curated PRS(24) for 11 HF risk factors (atrial arrhythmias, diabetes, heavy alcohol intake, hypertension, myocardial infarction, obesity, severe anaemia, smoking, stable angina, thyroid disorders and unstable angina(**Web Table 1**)), calculated for all UKB individuals with HF using PLINK 2.0(25). We assessed association with 12 HF SNPs(26) by extracting allelic dosages, inverted prior to analysis to reflect HF risk-increasing alleles. To test associations between PRS, HF SNPs and predicted clusters, we transformed predicted HF subtypes into 5 binary outcomes (cases: within cluster; controls: all other participants). By multiple logistic regression, we determined associations between HF-related PRS, SNPs and subtypes, visualising by heatmaps of p-values.

### (2) To propose pathways to improve impact (Impact)

#### Clinical utility

We assessed improved outcome prediction and open methods. We asked HF clinicians(n=5) about clinical relevance, justification and interpretability of results.

#### Effectiveness

We could not implement the large models. Therefore, based on clinician input, we developed: (i)a model (predicting cluster and survival using labels for identified clusters and clinically available factors, n=22), and (ii)a clinically usable R Shiny app which could be evaluated and used to evaluate effectiveness and cost-effectiveness in future studies. Analyses and visualisations were in R version 3.4.3, Python V3 and R Shiny.

#### Ethical approval

Approvals were by: (i) MHRA Independent Scientific Advisory Committee [18_217R]: Section 251 (NHS Social Care Act 2006), (ii) Scientific Review Committee [17THIN038-A1] and (iii) UKB 15422: Patient informed consent was not required or provided.

#### Data availability

All data produced in the present work are contained in the manuscript

## Results

### Development

#### Clinical relevance and Patients

We included 188,799, 124,263 (**Web Figures 2, 3**) and 9,573 individuals with incident HF in CPRD, THIN and UKB, respectively.

#### Algorithm

We selected 87/645 available factors after dimensionality reduction (**Web Figure 4, Web Table 2**).

### Validation

#### Internal

The optimal number of clusters was 5. Identified clusters were stable (Rand index, Jaccard means, and Fowlkes and Mallows’s indices >0.8 for all subtypes, using all ML methods except hierarchical clustering). Across datasets, we used similarity matrices to find the most representative algorithm, which was K-means (**Web figure 5**).

#### External

Subtypes were similar across datasets (c-statistic: 0.94, 0.80, 0.79, 0.83, 0.92 for the THIN model in CPRD and 0.79, 0.92, 0.90, 0.89, 0.92 for the CPRD model in THIN for subtypes 1-5, respectively)(**Web Table 3**).

Five clusters were identified based on demography, CVD risk factor burden, AF, CVD (particularly atherosclerotic disease), medications and laboratory factors. In CPRD, THIN and UK Biobank, we labelled the clusters as subtypes after studying each cluster’s characteristics: (1) Early-onset; (2) Late-onset; (3) AF-related; (4) Metabolic; and (5) Cardiometabolic. Distribution of subtypes was similar across THIN and CPRD, with Late-onset (30.9%) and Cardiometabolic (29.7%) the commonest and AF-related (8.9%) the least common(**Figure 2**).

**Figure 2.**
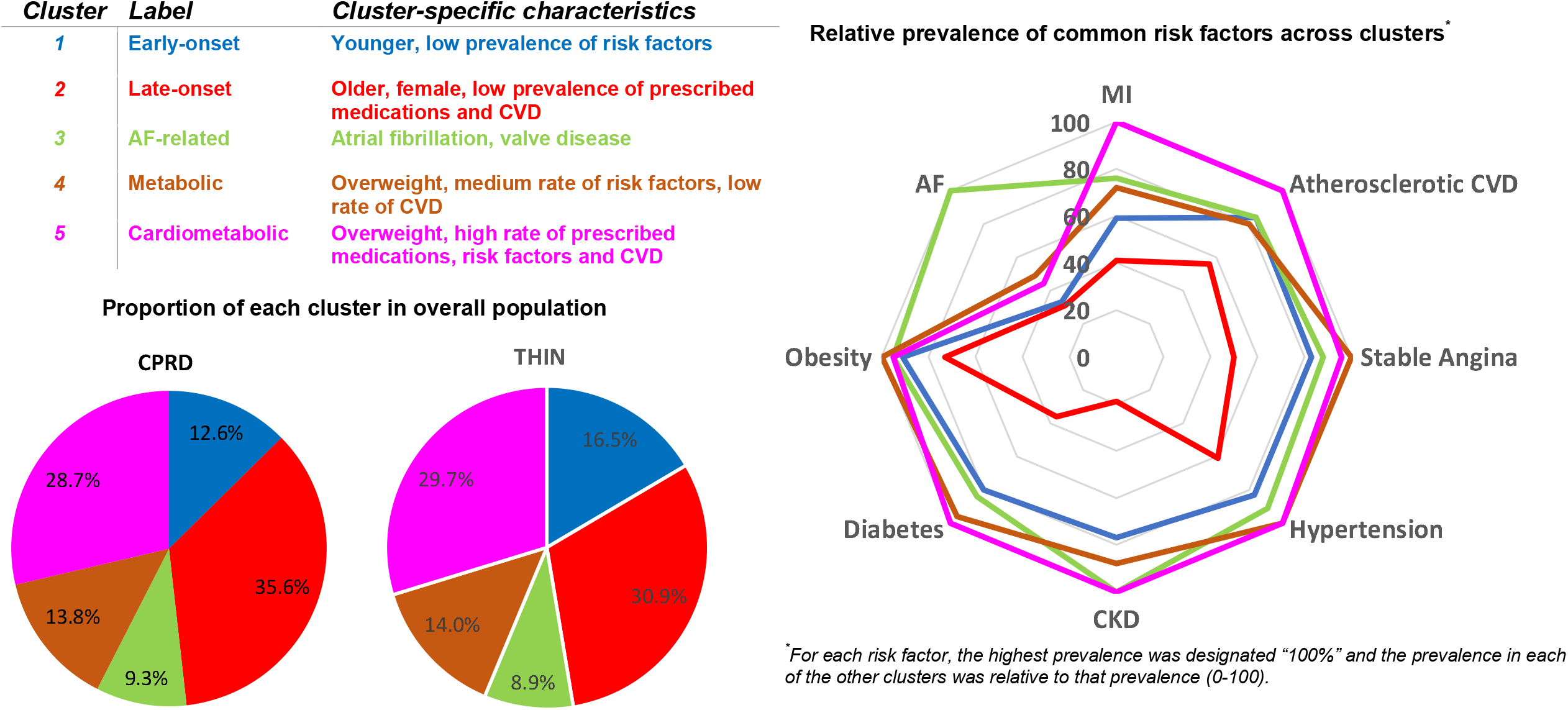
Externally validated clusters in incident heart failure in two UK primary care populations (n=313 062 overall; THIN and CPRD) *Abbreviations: AF:atrial fibrillation: CKD: chronic kidney disease; CPRD: Clinical Practice Research Datalink; CVD: cardiovascular disease; MI: myocardial infarction; THIN: The Health Innovation Network*.

Age and gender varied across subtypes (oldest: Late-onset; youngest: Early-onset; most females: Metabolic; and least females: Cardiometabolic). Prevalence of cardiovascular risk factors and diseases was highest in the Cardiometabolic subtype, e.g. hypertension 72.9%, obesity 34.3%, diabetes 41.1% and Atherosclerotic CVD 59.2% (**Table 1, Figure 2**). Age, blood investigations, BMI and blood pressure did not discriminate well between subtypes or mortality by subtype (**Table 1, Web Figures 6 and 7**).

**Table 1:** Baseline characteristics by subtype of incident heart failure in two UK primary care populations (n=313 062)

| **Subtypes**<br>Total N=313,062 | **1 (Early-onset)**<br>n=44,292 |  | **2 (Late-onset)**<br>n=105,610 |  | **3 (AF-related)**<br>n=28,617 |  | **4 (Metabolic)**<br>n=43,451 |  | **5 (Cardiometabolic)**<br>n=91,092 |  |
| --- | --- | --- | --- | --- | --- | --- | --- | --- | --- | --- |
| *Dataset* | **THIN**<br>n=20,503 | **CPRD**<br>n=23,789 | **THIN**<br>n=38,397 | **CPRD**<br>n=67,213 | **THIN**<br>n=11,059 | **CPRD**<br>n=17,558 | **THIN**<br>n=17,397 | **CPRD**<br>n=26,054 | **THIN**<br>n=36,906 | **CPRD**<br>n=54,186 |
| **Demographics** |  |  |  |  |  |  |  |  |  |  |
| Age-years (s.d.) | 69.1<br>(13.3) | 70.4<br>(12.4) | 79.9<br>(11.0) | 79.6<br>(12.3) | 73.6<br>(10.7) | 77.1<br>(10.5) | 74.0<br>(12.0) | 77.0<br>(11.3) | 75.3<br>(10.7) | 78.4<br>(10.5) |
| Female | 44.2 | 44.2 | 57.5 | 54.7 | 44.7 | 51.8 | 44.3 | 48.8 | 43.5 | 47.9 |
| **Risk Factors** |  |  |  |  |  |  |  |  |  |  |
| Obesity (BMI≥40) % | 3.6 | 3.4 | 1.5 | 2.0 | 5.2 | 3.5 | 5.0 | 5.0 | 4.7 | 4.8 |
| BMI-kg/m<sup>2</sup> (s.d.) | 27.7<br>(5.9) | 27.9<br>(5.7) | 25.5<br>(5.0) | 26.2<br>(5.2) | 28.5<br>(6.3) | 27.7<br>(5.9) | 28.7<br>(6.3) | 28.5<br>(6.4) | 28.5<br>(6.2) | 28.1<br>(6.2) |
| Hypertension | 58.1 | 85.8 | 43.5 | 61.8 | 65.6 | 82.8 | 81.4 | 90.3 | 72.9 | 89.4 |
| Diastolic pressure | 77.1<br>(12.9) | 78.1<br>(13.0) | 77.3<br>(11.9) | 78.3<br>(11.4) | 78.2<br>(12.2) | 77.3<br>(12.2) | 74.9<br>(11.8) | 74.6<br>(11.7) | 74.3<br>(11.8) | 73.7<br>(11.6) |
| Systolic pressure | 133.5<br>(22.9) | 136.9<br>(13.0) | 138.4<br>(22.6) | 139.8<br>(11.4) | 137.7<br>(22.5) | 139.6<br>(12.2) | 131.6<br>(19.9) | 132.9<br>(11.7) | 132.1<br>(21.1) | 133.8<br>(11.6) |
| Total Cholesterol-<br>mmol/L (s.d.) | 4.8<br>(1.3) | - | 5.1<br>(0.83) | - | 4.8<br>(1.1) | - | 4.4<br>(1.1) | - | 4.4<br>(1.2) | - |
| Diabetes % | 26.3 | 35.7 | 10.1 | 16.7 | 27.7 | 30.5 | 31.1 | 37.9 | 34.5 | 40.1 |
| CKD Stage 3 | 30.8 | 33.3 | 7.5 | 5.3 | 36.0 | 26.9 | 36.0 | 35.4 | 39.4 | 34.2 |
| Renal failure | 0.76 | 1.1 | 0.36 | 0.27 | 0.73 | 1.1 | 0.43 | 0.98 | 1.1 | 0.82 |
| Creatinine-<br>mmol/L (s.d.) | 104.4<br>(48.7) | 106.6<br>(49.8) | 115.3<br>(57.3) | 107.0<br>(60.8) | 107.5<br>(45.8) | 109.5<br>(57.8) | 103.3<br>(50.1) | 104.3<br>(61.2) | 112.9<br>(61.5) | 112.5<br>(65.2) |
| Haemoglobin-<br>g/dL (s.d.) | 13.3<br>(1.8) | 13.2<br>(1.9) | 12.6<br>(1.6) | 12.9<br>(1.7) | 13.1<br>(1.8) | 12.9<br>(1.8) | 13.1<br>(1.8) | 12.9<br>(1.9) | 12.9<br>(1.8) | 12.8<br>(1.9) |
| **BASELINE CVD** |  |  |  |  |  |  |  |  |  |  |
| AF | 31.9 | 65.0 | 28.2 | 45.0 | 93.3 | 98.6 | 46.1 | 65.6 | 40.1 | 58.3 |
| MI | 32.1 | 46.6 | 15.1 | 23.1 | 28.2 | 40.2 | 27.4 | 37.7 | 37.8 | 44.3 |
| Unstable Angina | 6.6 | 21.6 | 3.2 | 7.2 | 7.5 | 20.7 | 7.3 | 18.6 | 11.6 | 20.5 |
| Stable Angina | 60.8 | 73.2 | 39.0 | 43.5 | 63.4 | 69.6 | 73.5 | 74.0 | 72.9 | 73.3 |
| Stroke | 15.8 | 19.7 | 14.4 | 15.4 | 19.0 | 20.4 | 16.8 | 16.8 | 20.1 | 21.9 |
| Peripheral<br>Vascular Disease | 13.7 | 23.4 | 11.8 | 15.5 | 16.1 | 27.4 | 14.8 | 24.6 | 20.1 | 27.5 |
| Atherosclerotic<br>CVD | 49.0 | 64.8 | 34.5 | 43.3 | 49.1 | 63.8 | 46.6 | 57.8 | 59.2 | 66.3 |
| **Drugs at**<br>**Baseline** |  |  |  |  |  |  |  |  |  |  |
| Antiplatelet | 74.9 | 80.3 | 54.2 | 44.8 | 80.0 | 76.5 | 77.8 | 77.8 | 88.7 | 86.0 |
| Statin | 74.8 | 79.4 | 12.7 | 8.2 | 66.1 | 53.5 | 76.5 | 74.7 | 88.7 | 88.5 |
| ACEI | 84.3 | 89.3 | 52.6 | 50.4 | 86.5 | 83.8 | 86.9 | 88.7 | 91.7 | 85.3 |
| Beta blocker | 73.6 | 71.4 | 28.9 | 29.3 | 73.7 | 57.6 | 87.1 | 79.3 | 81.2 | 70.1 |
| Diuretics | 88.1 | 88.5 | 88.5 | 71.7 | 96.0 | 91.0 | 90.1 | 87.4 | 93.6 | 87.4 |
| Aldosterone<br>Antagonist | 38.3 | - | 17.4 | - | 47.5 | - | 40.7 | - | 38.6 | - |
| Warfarin | 29.9 | 52.4 | 18.0 | 27.4 | 60.9 | 50.5 | 39.5 | 46.3 | 35.8 | 35.7 |

ACEI: angiotensin converting enzyme inhibitor; AF: atrial fibrillation; BMI: body-mass index; CKD: chronic kidney disease; CPRD: Clinical Practice Research Datalink; CVD: cardiovascular disease; THIN: The Health Innovation Network.

#### Prognostic

In CPRD using the THIN model, 1-year mortality was 2%, 46%, 6%, 11% and 37% for subtypes 1 to 5 respectively, with c-statistic of 0.68, 0.62, 0.57, 0.71 and 0.68. There were differences in mortality for clusters 2 and 5 respectively, but not other clusters, between THIN and CPRD (**Figure 3, Web Table 6**). AF occurred after HF diagnosis in the AF-related subtype and was more likely to be before HF diagnosis for other subtypes. Hypertension, myocardial infarction, stroke and peripheral vascular disease occurred predominantly before HF diagnosis in the Cardiometabolic subtype, and after HF diagnosis in AF-related and Early-onset subtypes (**Figure 4, Web figure 8**). After HF diagnosis, use of beta-blockers, ACEI and aldosterone antagonists was highest in those with AF-related and Early-onset subtypes (**Web Figure 9**).

**Figure 3.**
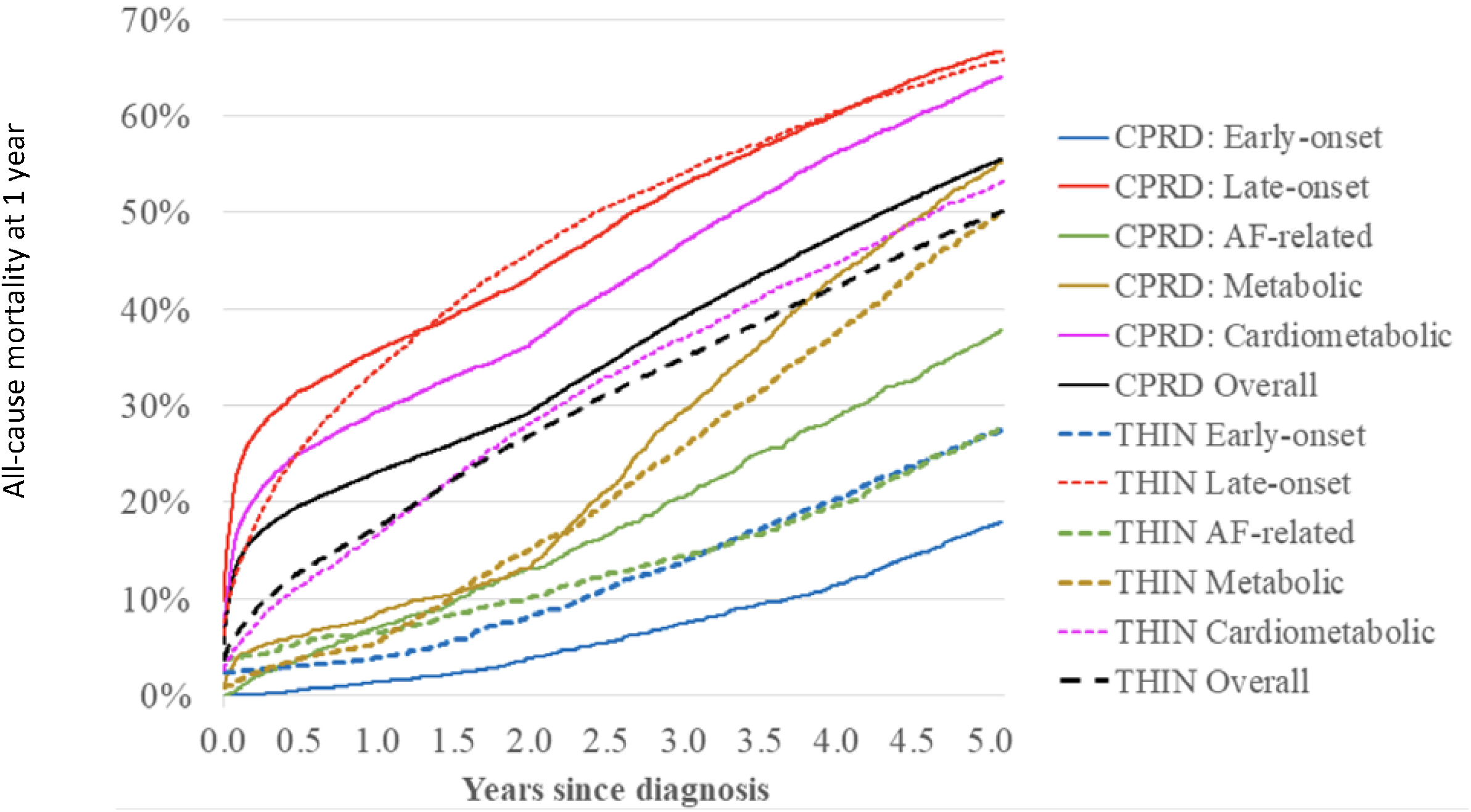
Prognostic validation using clusters for all-cause mortality in incident heart failure in two UK primary care populations (n=313 062 overall; THIN and CPRD) *\*for pairwise comparisons using Log-rank test, please see* ***Web Table 6***

**Figure 4.**
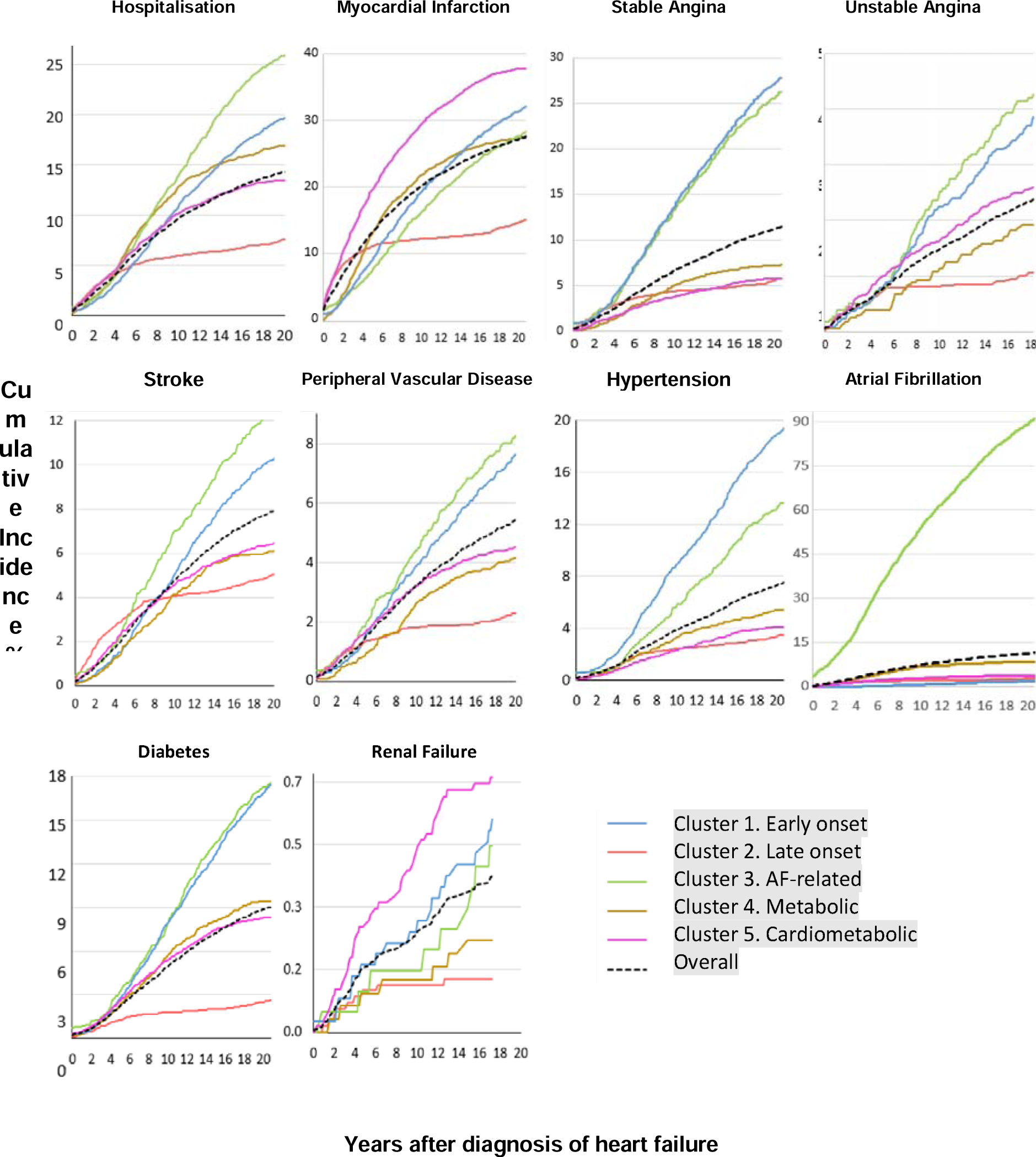
Risk of non-fatal cardiovascular diseases and all-cause hospitalisation in five heart failure subtypes after diagnosis.

#### Genetic

7801 out of 9573 individuals in UKB had necessary genetic data for analyses of PRS and SNPs. Numbers in the “Metabolic” cluster were low (n=49), but other clusters were well-represented (Late-onset: 1586; AF: 1981; Early-onset: 1553; Cardiometabolic: 2633).

The associations between HF subtypes and both PRS and HF-related SNPs are depicted in **Figure 5** (see also Web **Table 5**). PRS for Atrial Arrhythmias, Diabetes, Hypertension, Myocardial Infarction, Obesity, Stable and Unstable Angina were all associated with one or more HF subtypes after correction for multiple testing (p-value < 9.09 × 10^−4^). Late-onset and Cardiometabolic HF subtypes broadly associated with similar PRS. No PRS was associated with the Metabolic subtype, potentially due to small numbers in this group (n=49). Eight SNPs were nominally associated (p=0.05) with predicted HF subtypes. Four of these associated SNPs were confined to the AF-related subtype: rs11745324, rs17042102, rs4746140 and rs4766578 (**Figure 5b**), corresponding to the PITX2/FAM241A, SYNPO2L, AGAP5 and ATXN2 gene regions respectively. Associations between rs17042102 and AF-related subtype persisted even after correcting for multiple testing (0.05/60 = 8.3×10^−4^), suggesting importance of chromosome 4 in AF-related HF.

**Figure 5.**
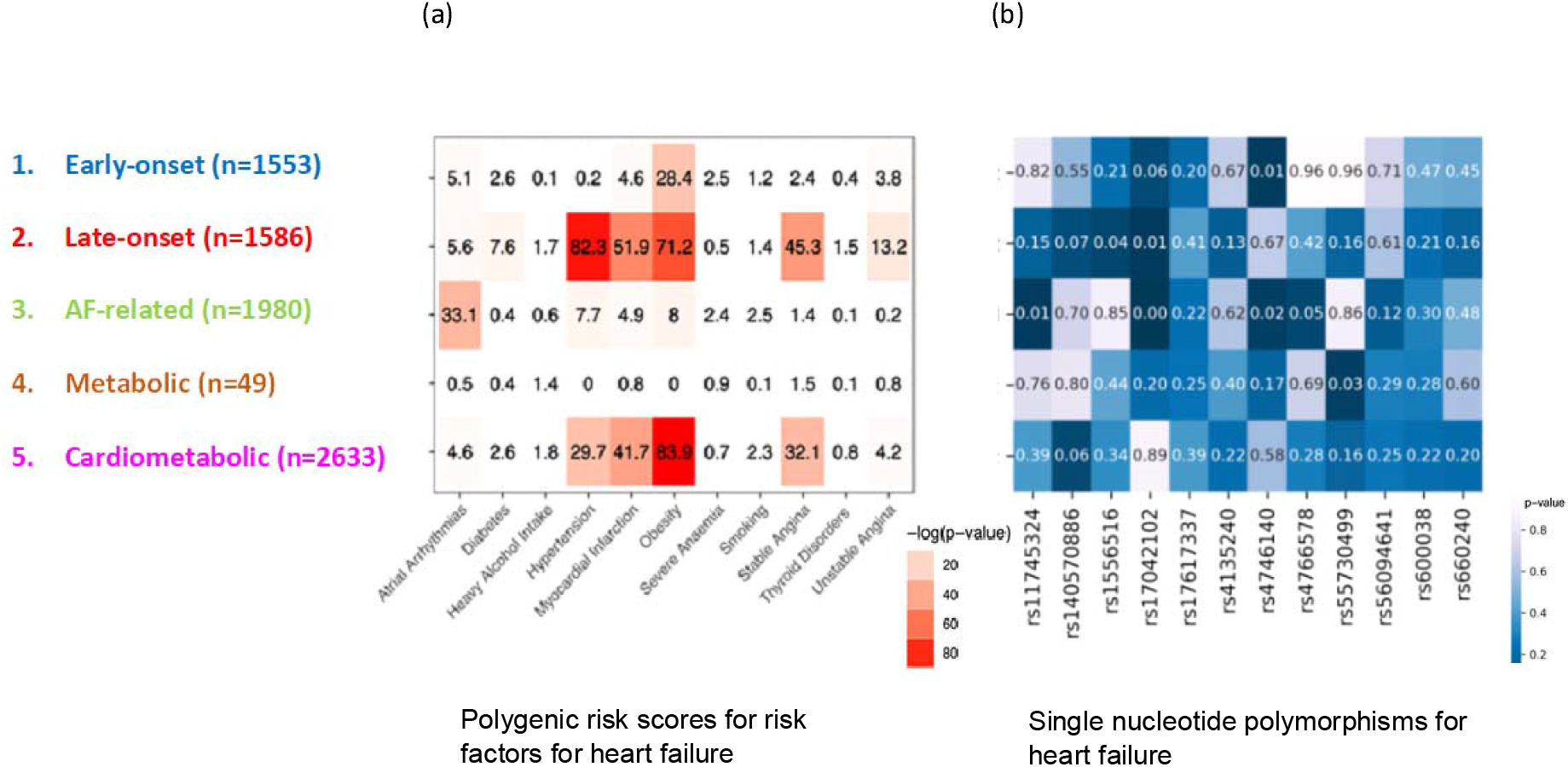
Heart failure subtypes and genotype associations (n = 7801): (a) association between heart failure subtypes and polygenic risk scores for 11 related traits; (b) direct association between heart failure subtypes and 12 related single nucleotide polymorphisms. *Numbers represent p-values from the logistic regression of (a) polygenic risk score or (b) single nucleotide polymorphism versus cluster output*.

### Impact

#### Clinical utility

Sample clinicians (n=5) reported that the included factors and the identified clusters had clinical relevance. Differences between clusters by baseline characteristics and survival were distinguishable and interpretable. The framework/methods proposed were transparent and generalisable.

#### Effectiveness

We developed an open access app which can be used by clinicians to identify the cluster which a particular patient falls within, and their predicted survival (https://pasea.shinyapps.io/hf_cluster_app/). The interviewed clinicians felt that this app was a feasible use of the identified clusters in care and that it could enable testing of effectiveness and cost effectiveness (as elaborated in the discussion).

Based on clinician and researcher input, effectiveness could be tested in six ways: (i)prospective validation of the clusters in care; (ii)comparison of treatment and care pathways with app for cluster identification versus usual care (possibly in a trial); (iii)predictive accuracy for survival compared with existing risk prediction tools; (iv) patient-reported outcomes; (v)patient satisfaction; and (vi)clinician satisfaction and ease of use in clinical practice. Cost effectiveness could be estimated by modelling the impact on care and outcomes based on the above analysis of effectiveness, the time required to estimate and communicate subtype in clinical settings, and the potential effect on healthcare utilisation and outcomes.

## Discussion

To our knowledge, this is the first study to define and validate data-driven heart failure disease clusters across multiple ML methods, nationally representative datasets and multiple validation methods. We make three distinct advances. First, we identified five incident HF subtypes: Early-onset; Late-onset; AF-related; Metabolic; and Cardiometabolic. Second, we confirmed internal, external, prognostic and genetic validity. Third, we developed a means of using identified subtypes in clinical practice and suggest ways of evaluating effectiveness.

Our five subtypes are compatible with two major clustering studies(12, 13). The Cardiometabolic subtype could represent ischaemic aetiology(1, 2, 9). For aetiology, guidelines and research, studies have predominantly focused on ischaemic versus non-ischaemic HF, and HF defined by cut-offs based on EF(27). The AF-related subtype is consistent with doubling of risk of incident AF observed in prevalent HF, compared to no HF (HR 2.18; 95% CI, 1.26–3.76)(28). The high proportion of prevalent AF in other subtypes is consistent with “AF begets HF”. Further study of atrial, ventricular and atrio-ventricular cardiomyopathies will inform temporal associations between AF and different HF subtypes(29). Individuals in the Metabolic subtype were younger with higher prevalence of AF and obesity(30), but lower prevalence of atherosclerotic CVD, than the Cardiometabolic subtype, though not entirely distinguishable. Age is predictive of overall HF and particular subtypes in prior studies. Therefore, Early-onset and Late-onset subtypes are plausible(12), warranting further investigation across countries and factors, e.g. echocardiography.

Methodologically, we offer advances in external validation of ML in subtype classification and in risk prediction in HF, which has been rare (only 4/27 and 2/31 studies respectively) and in small samples (n=44 to 3203 for studies of HF subtypes)(13). Our subtypes showed good accuracy within and across datasets, and good predictive accuracy for Early-onset, Metabolic and Cardiometabolic subtypes, though less accurate for AF-related and Late-onset subtypes. The c-statistic for LVEF, the most commonly used feature to define HF subtype, was only 0.52 in a large Swedish national registry study using ML(12). Even after inclusion of more clinical factors (e.g., echocardiography, NT-pro-BNP) or focus on certain subgroups or clinical scenarios(31), improved risk prediction for mortality and other outcomes remains challenging. Our finding of PRS and SNPs associated with the AF-related subtype are novel, signalling potential utility of assessment for biologic validity of cluster analyses and their linkage to EHR(26). The mild associations observed with related PRS for the Early onset HF subtype (with the exception of strong association with obesity and atrial arrhythmias), compared to Late-onset and Cardiometabolic subtypes, are of interest. Studies of ML in HF should focus on further validation in representative datasets from other countries, disease definition and use of high-dimensionality proteomics and imaging data.

Recent guidelines describe the need for systematic approaches to design, evaluation and implementation of ML in healthcare(13, 32, 33). We address issues at the development and validation stages to use of ML for subtype classification and risk prediction in HF. Our robust, structured framework of internal, external, prognostic and genetic validation could extend acceptability and generalisability of ML to clinical practice and is transferable to other diseases. Our approach to clinical utility (relevance, justification, and interpretability) illustrates how specialist and patient views can be assessed and incorporated in the evaluation of ML in healthcare, where there is currently little guidance to aid implementation in healthcare. Although we interviewed a limited number of clinicians, the approach could be used at national and international level. To assess effectiveness, we offer a prototype for application of our identified subtypes in care which needs further investigation at the implementation stage, especially analyses of effectiveness and cost-effectiveness, which are currently lacking.

This is one of the largest EHR analysis to-date to use ML in subtype classification and risk prediction of HF, and for the first time, investigating multiple ML methods, multiple nationally representative datasets and multiple validation methods. By using EHR, our derivation and validation cohorts are representative of real-world patients, increasing confidence that evaluation in clinical practice is worthwhile. We incorporated factors before and after HF diagnosis, enabling insights into trajectory as well as aetiology. However, there are several limitations. First, we are using EHR phenotypes of HF, which do not have complete biochemical (e.g. NT-pro-BNP) and imaging (e.g. LVEF) profiles, and therefore certain previous classifications are not possible (e.g. HF with preserved ejection fraction). However, NT-pro-BNP and LVEF tend to be more available in secondary and tertiary settings and are commonly not available in unselected patients with new HF, where 26% were never hospitalised(19). Moreover, our phenotypes have been validated and used in prior large-scale studies(19). Second, although we used 645 factors, risk factor phenotypes are limited by timing and accuracy of clinician recording in the EHR, which may affect analyses of factors before and after HF. Third, although we use two large, nationally representative primary care datasets, they are both from the UK and may not be representative of HF in other countries or settings. Fourth, we performed only supervised analyses of PRS of 11 traits related to HF and 12 SNPs previously associated with HF (and numbers in the Metabolic cluster were small), necessitating further genetic analysis in larger cohorts.

## Conclusions

Across three large, population-scale datasets, four machine learning methods, 645 factors, and four validation methods, we identify five heart failure subtypes with good discriminatory accuracy within and across datasets, and good predictive accuracy for 1-year mortality. These subtypes may have implications for research in terms of use of EHR and ML to identify HF subtypes in future clinical trials and observational studies, as well as clinical practice in terms of management and prognosis.

## Supporting information

Web Table 1

## Data Availability

All data produced in the present work are contained in the manuscript

## Contributors

AB and HH conceived the research question. AB, SC, MD, GF, SD and HH designed the study and analysis plan. SC, MD, LP, JHT and GF conducted different parts of experiments. AB, MD, SC and HH drafted the initial and final versions of manuscript. All authors critically reviewed early and final versions of the manuscript.

## Declaration of interests

AB is supported by research funding from the National Institute for Health Research (NIHR), British Medical Association, AstraZeneca, and UK Research and Innovation. BT and TD are employees of Bayer. All other authors declare no competing interests.

## Acknowledgments

HH is supported by Health Data Research UK (grant No. LOND1), which is funded by the UK Medical Research Council, Engineering and Physical Sciences Research Council, Economic and Social Research Council, Department of Health and Social Care (England), Chief Scientist Office of the Scottish Government Health and Social Care Directorates, Health and Social Care Research and Development Division (Welsh Government), Public Health Agency (Northern Ireland), British Heart Foundation, and Wellcome Trust. HH is a NIHR Senior Investigator. AB, SD, FA and HH are funded by the NIHR University College London Hospitals Biomedical Research Centre. All authors are supported by the BigData@Heart Consortium, funded by the Innovative Medicines Initiative-2 joint undertaking under grant agreement no 116074. This joint undertaking receives support from the EU’s Horizon 2020 research and innovation programme and EFPIA.

