## Supplementary methods, tables and figures for "Identifying subtypes of heart failure with machine learning: external, prognostic and genetic validation in three electronic health record sources with 320,863 individuals"

**Web Appendix**

| **Contents** | **Page number** |
| --- | --- |
| Web Figure 1 Deduplication algorithm for overlap between THIN and CPRD | 2 |
| Web Figure 2 Source of heart failure diagnoses in linked primary and secondary care data (CPRD, HES, ONS) | 3 |
| Web Figure 3 Study population for subtyping analysis | 4 |
| Web figure 4. Variable importance for dimensionality reduction (n=87) | 5 |
| Web Figure 5. Comparison of Clustering Methods | 6 |
| Web figure 6. Selected continuous variables across five subtypes of individuals with incident heart failure. | 7 |
| Web figure 7. Mortality in subtypes of heart  failure by continuous variables | 8 |
| Web figure 8. Risk of non-fatal cardiovascular diseases and all-cause hospitalisation in five heart failure subtypes before or after diagnosis. | 9 |
| Web Figure 9. Rates of medication use in five heart failure subtypes ever or after diagnosis. | 10 |
| Web Table 1: Covariates included in clustering analyses (n=635)- separate Excel file | 11 |
| Web Table 2. Supplementary methods used for Heart failure subtype clustering. | 12 |
| Web Table 3 External validity: performance of four clustering methods in two UK primary care populations (CPRD and THIN) | 15 |
| Web Table 4. Heart failure single nucleotide polymorphisms (SNPs) included in analysis of biologic validity for subtypes of heart failure. | 16 |
| Web Table 5. Polygenic risk scores examined from the Polygenic Score Catalog. | 17 |
| Web Table 6. Pairwise comparisons (p-values) of survival probability of discovered subtypes in CPRD and THIN data using Log-rank test. | 18 |

**Web Figure 1 Deduplication algorithm for overlap between THIN and CPRD**

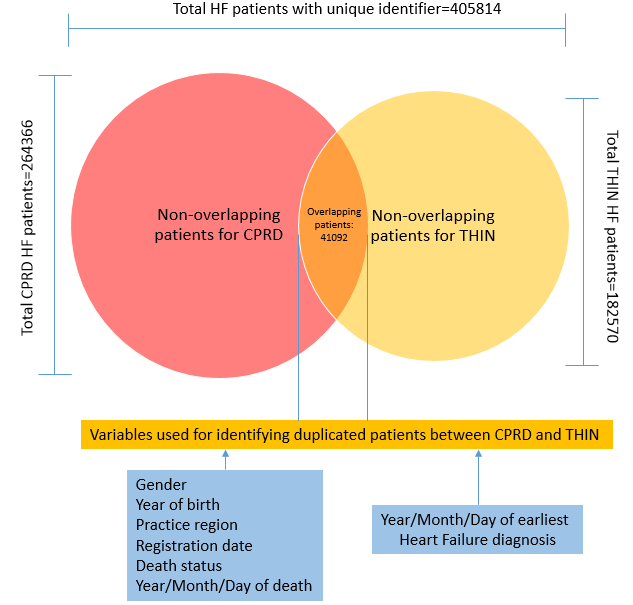

**Web Figure 2 Source of heart failure diagnoses in linked primary care (CPRD) and secondary care (HES) and cause of death (ONS) data.**

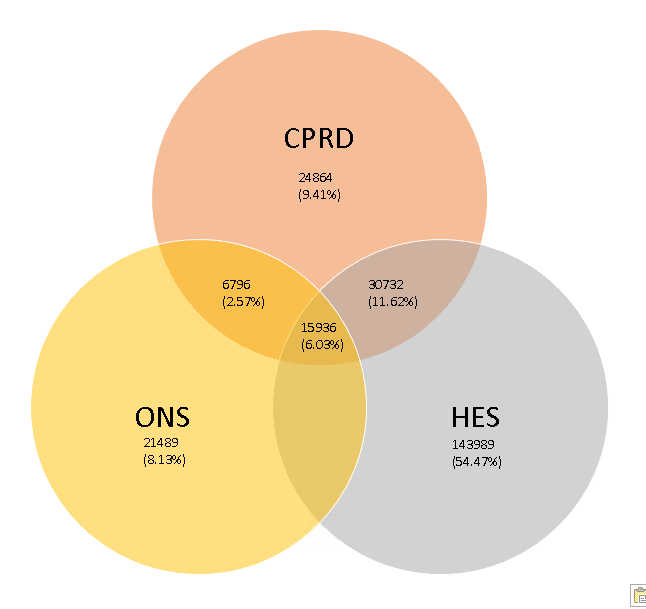

**Web Figure 3 Study population for subtyping analysis**

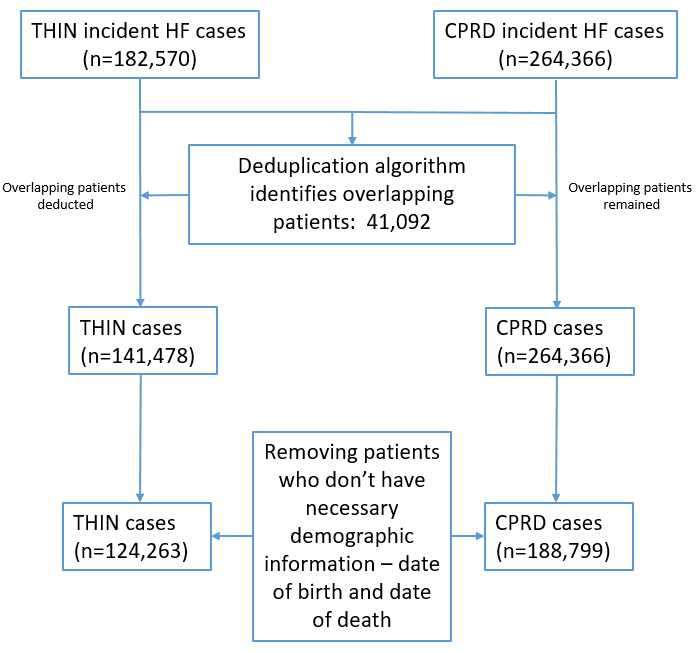

**Web figure 4. Variable importance for dimensionality reduction (n=87)**

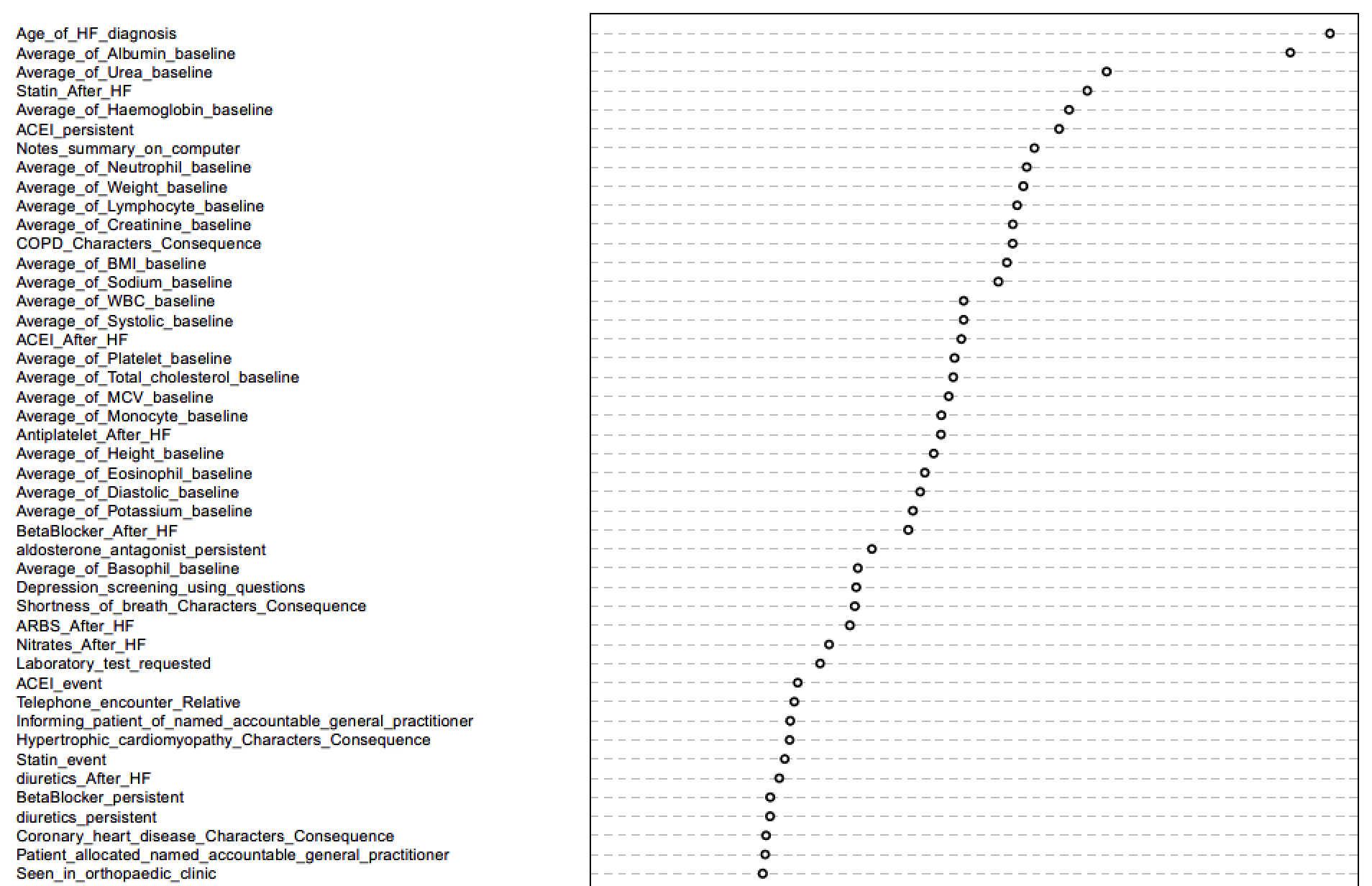

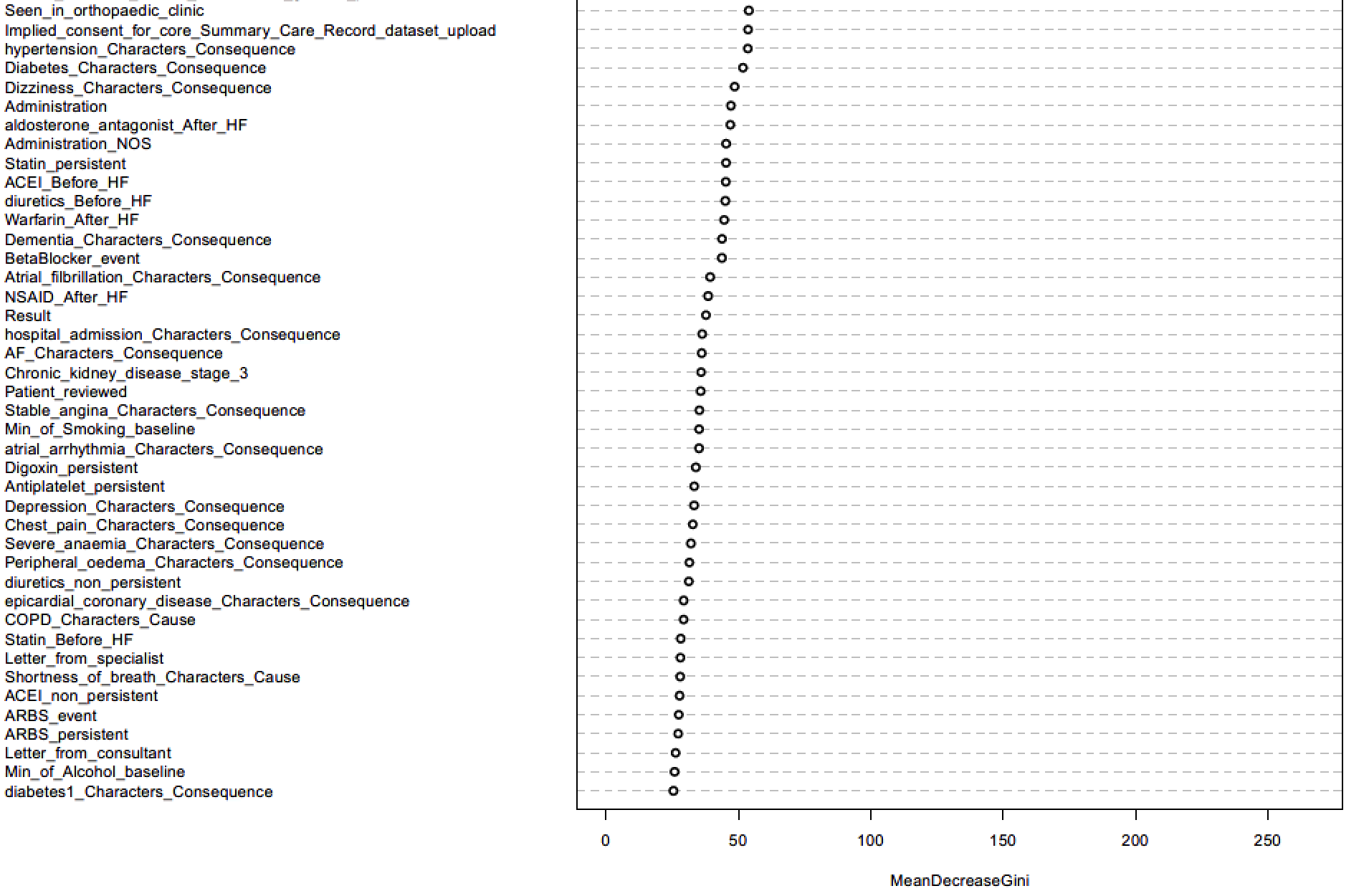

**Web Figure 5. Comparison of Clustering Methods**

1) **Cluster similarity metrics**

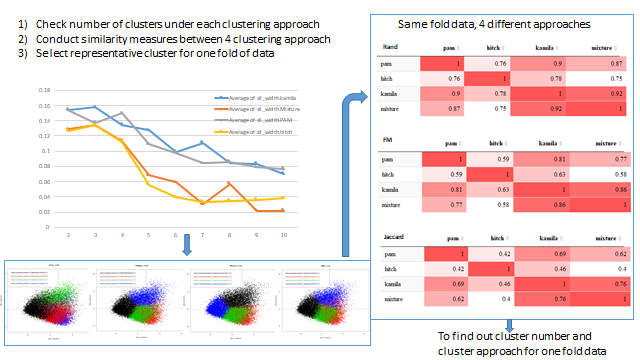

2) **Selection of representative clusters**

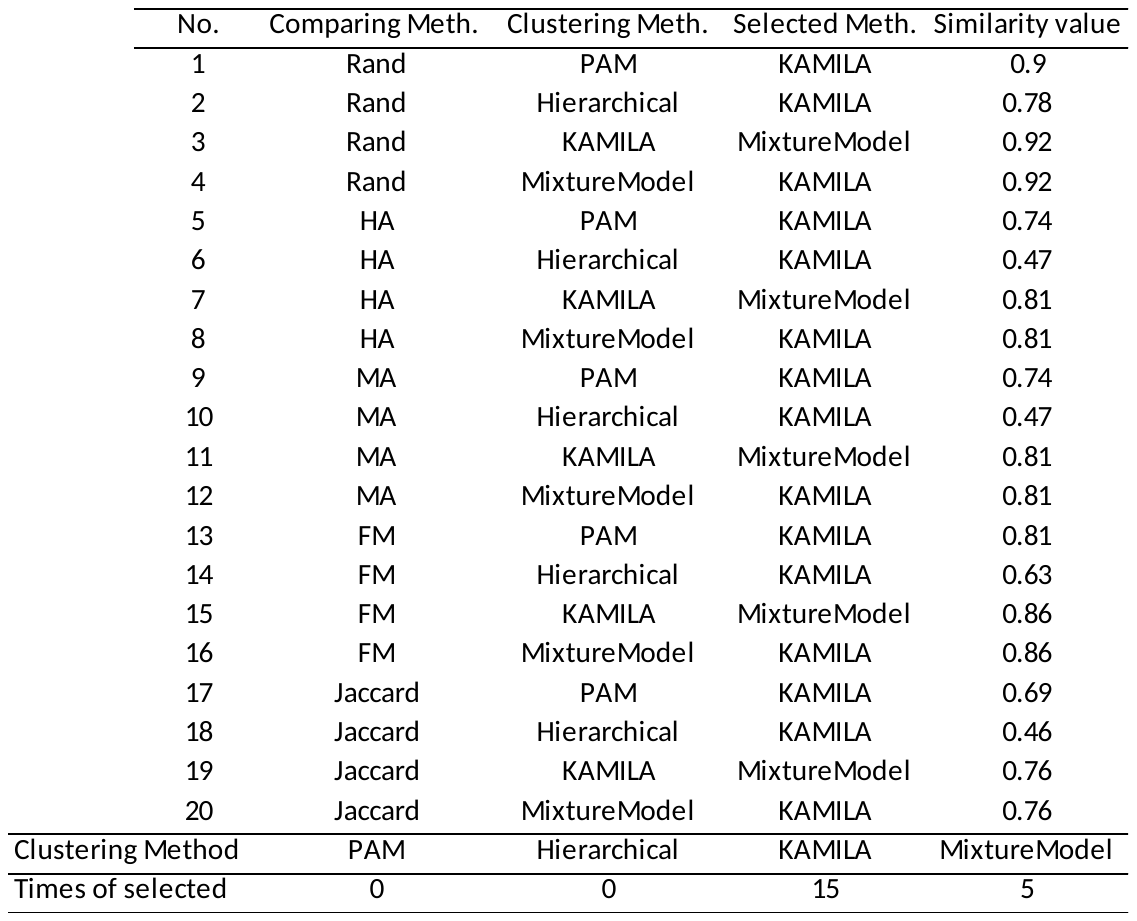

**Web figure 6. Selected continuous variables across five subtypes of individuals with incident heart failure.**

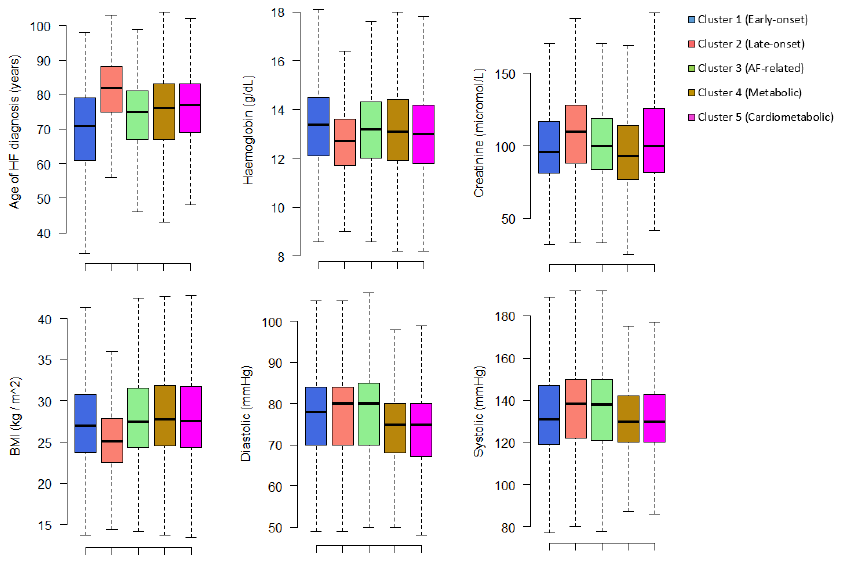

**Web figure 7. Mortality in subtypes of heart failure by continuous variables**

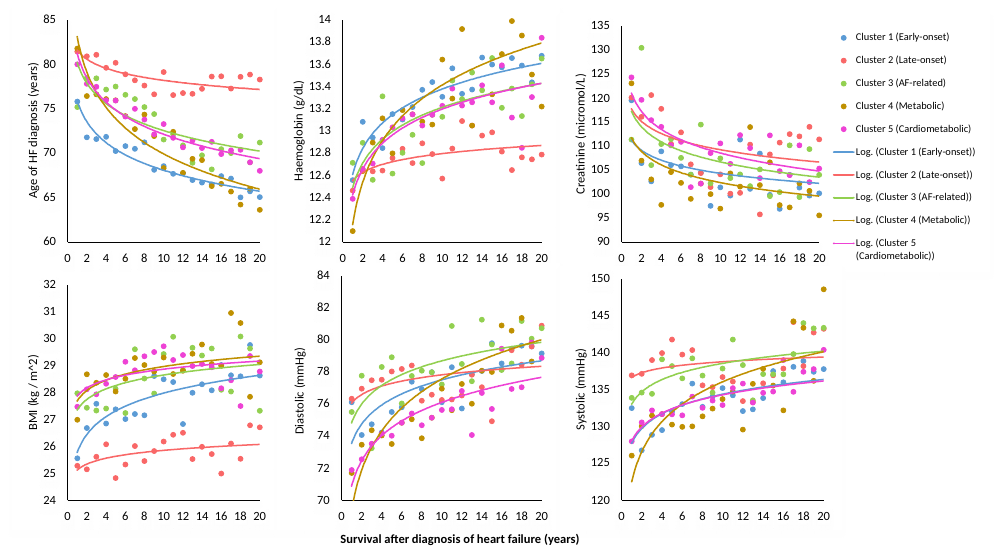

**Web figure 8. Risk of non-fatal cardiovascular diseases and all-cause hospitalisation in five heart failure subtypes before or after diagnosis.**

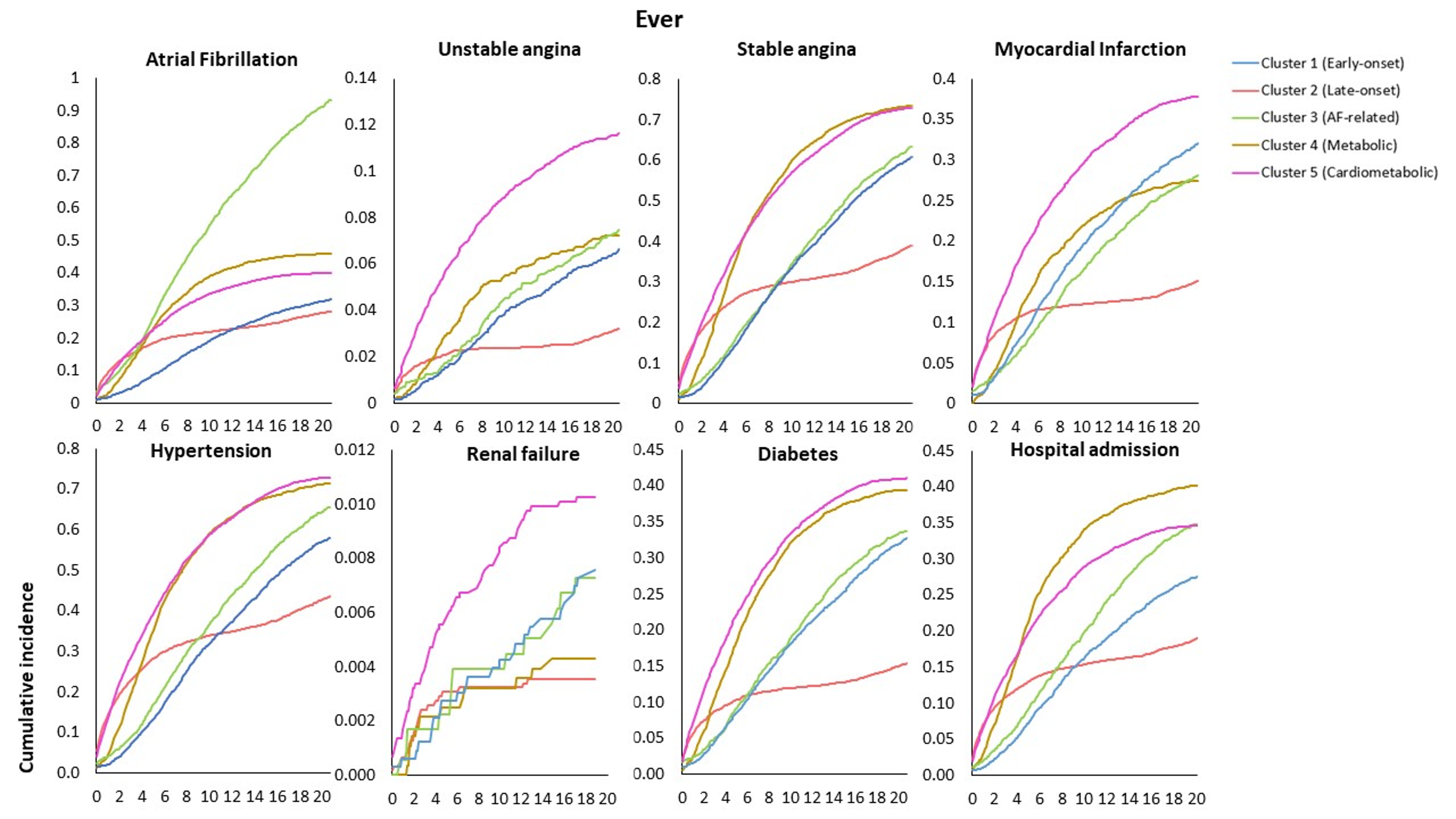

**Web Figure 9. Rates of medication use in five heart failure subtypes ever or after diagnosis.**

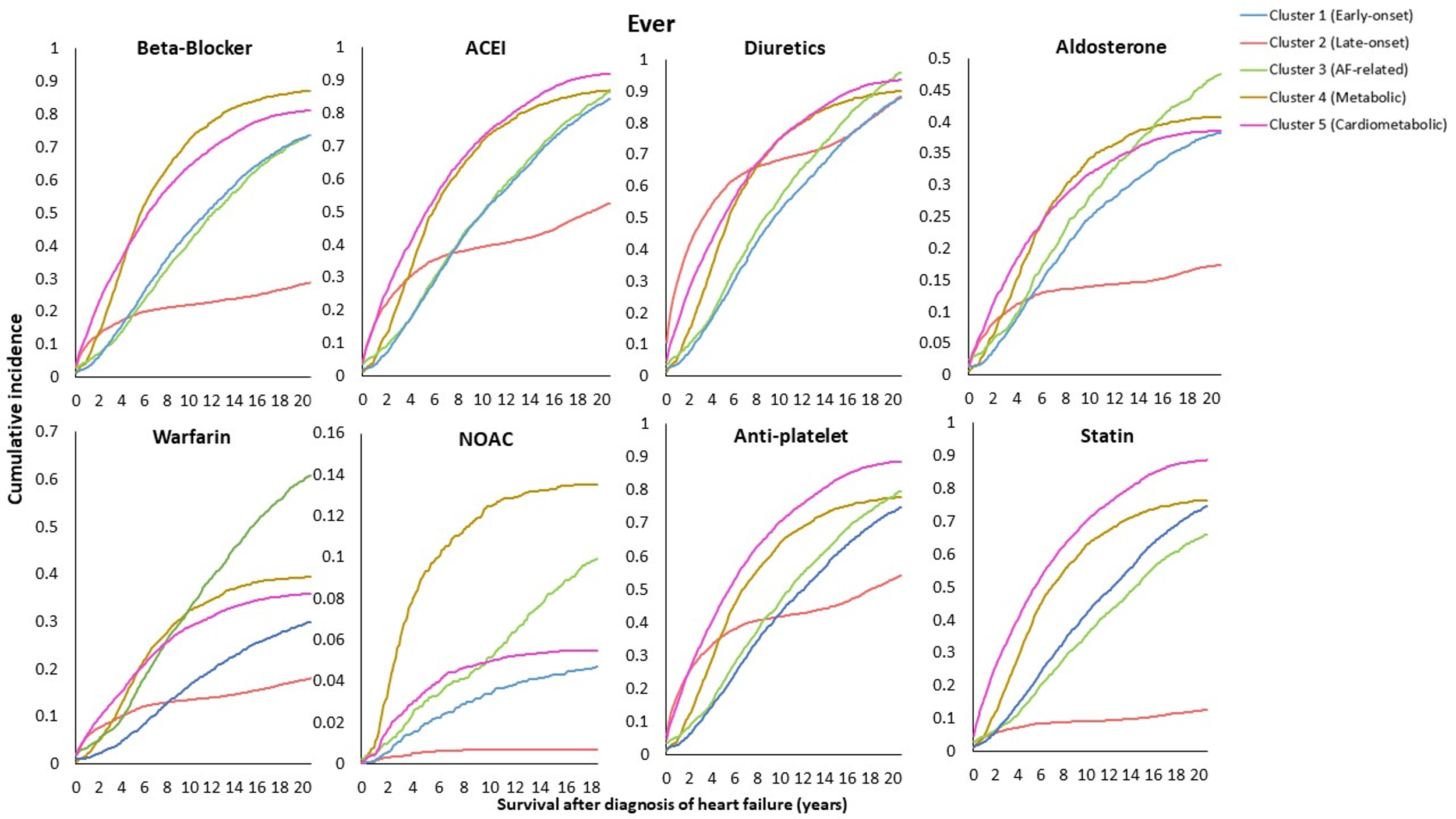

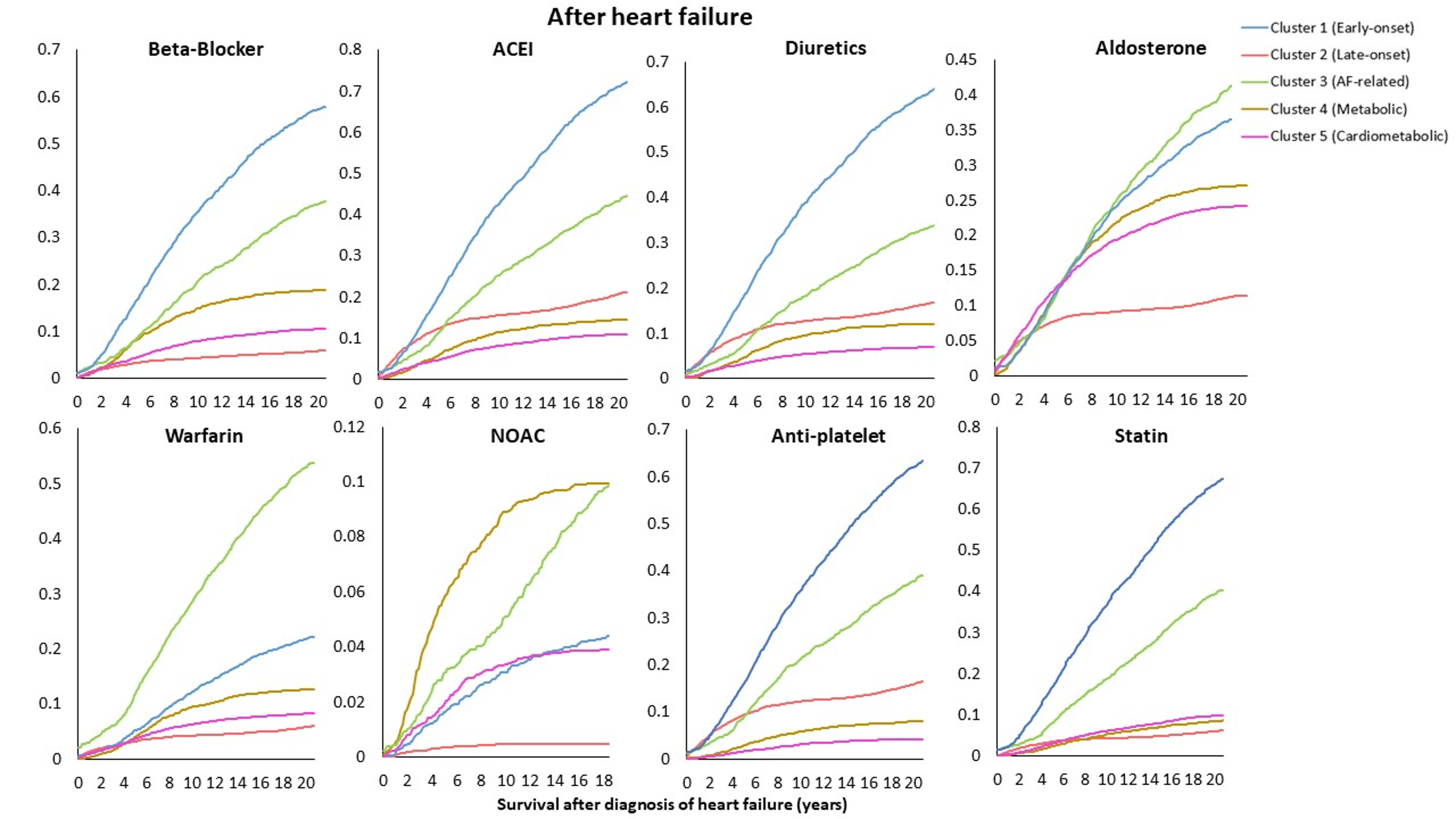

**Web Table 1: Covariates included in clustering analyses (n=635)**

**See Web Table 1 Excel attachment.**

**Web Table 2. Supplementary methods**
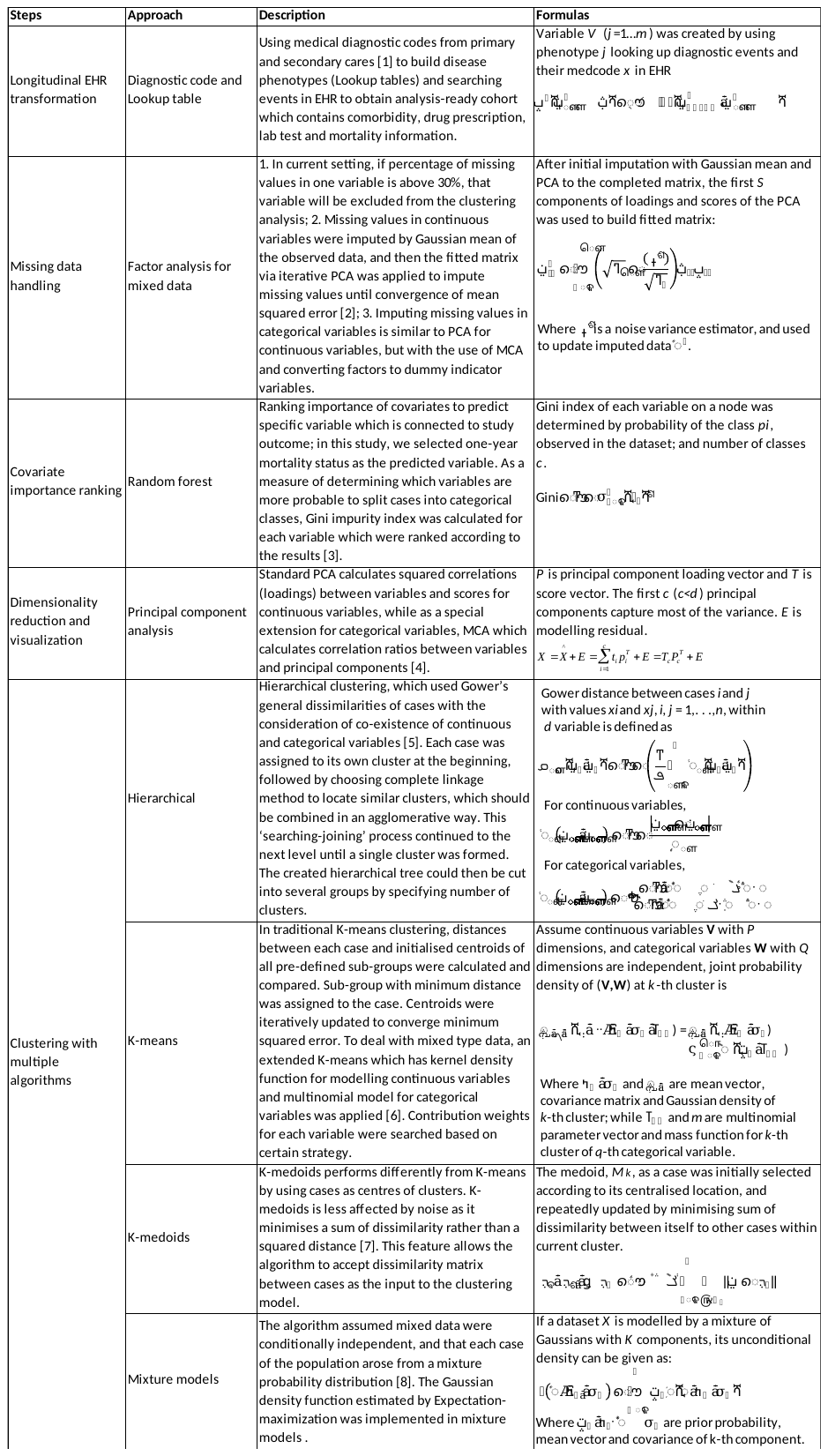

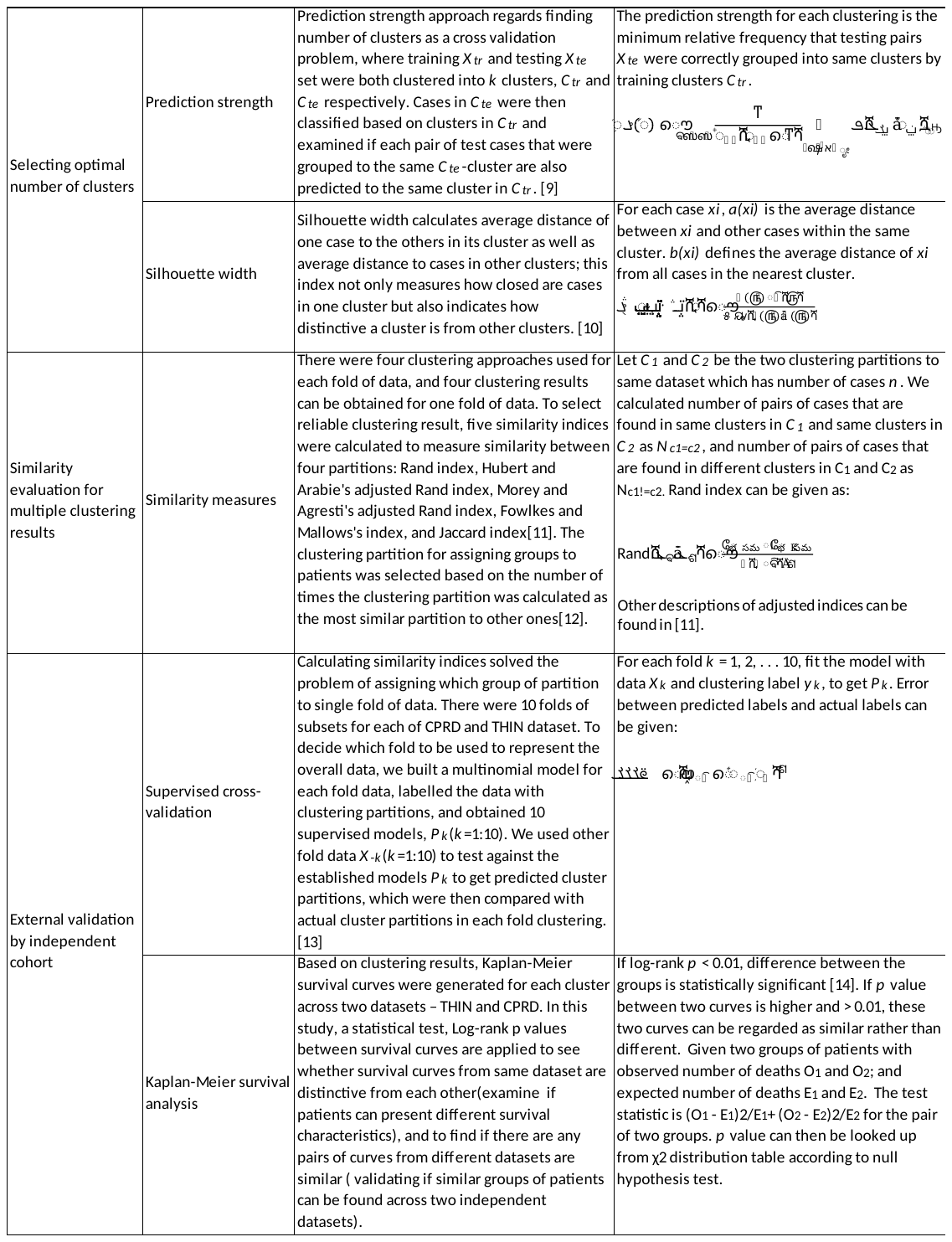

**Web Table 3. External validation: performance of four clustering methods in two UK primary care populations (CPRD and THIN)**

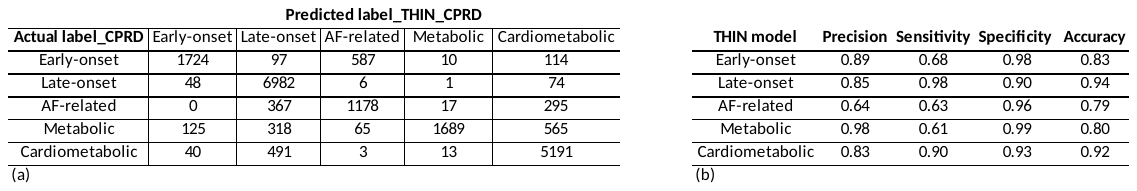

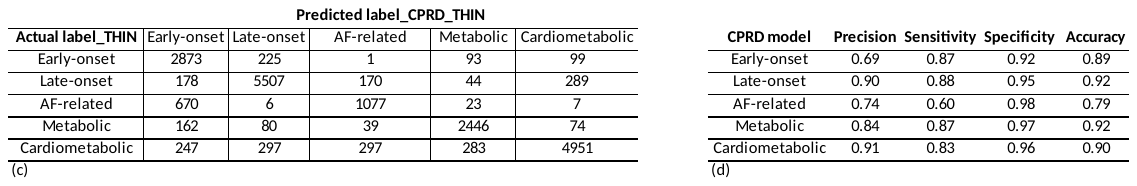

**Web Table 4. Heart failure single nucleotide polymorphisms (SNPs) included in analysis of biologic validity for subtypes of heart failure.**

| **HERMES** | | | | | | | | **UKB** | | |
| --- | --- | --- | --- | --- | --- | --- | --- | --- | --- | --- |
| **CHR** | **SNP** | **POS** | **Gene Region** | **effect allele** | **other allele** | **OR (95% CI)** | **P value** | **A1*** | **A2** | **Flip UKB Dosage** |
| 1 | rs660240 | 1.1E+08 | CELSR2 | C | T | 1.06 (1.04–1.08) | 3.25E-10 | T | C | TRUE |
| 4 | rs17042102 | 1.12E+08 | PITX2, FAM241A | A | G | 1.12 (1.09–1.14) | 5.71E-20 | G | A | TRUE |
| 5 | rs11745324 | 1.37E+08 | KLHL3 | G | A | 1.05 (1.03–1.07) | 2.35E-08 | G | A | FALSE |
| 6 | rs4135240 | 36647680 | CDKN1A | T | C | 1.05 (1.03–1.07) | 6.84E-09 | T | C | FALSE |
| 6 | rs55730499 | 1.61E+08 | LPA | T | C | 1.11 (1.08–1.14) | 1.83E-11 | C | T | TRUE |
| 6 | rs140570886 | 1.61E+08 | LPA | C | T | 1.24 (1.16–1.3) | 7.69E-11 | T | C | TRUE |
| 9 | rs1556516 | 22100176 | 9p21/CDKN2B-AS1 | C | G | 1.06 (1.05–1.08) | 1.57E-15 | G | C | TRUE |
| 9 | rs600038 | 1.36E+08 | ABO, SURF1 | C | T | 1.06 (1.04–1.08) | 3.68E-09 | T | C | TRUE |
| 10 | rs4746140 | 75417249 | SYNPO2L, AGAP5 | G | C | 1.07 (1.05–1.09) | 1.10E-09 | G | C | FALSE |
| 10 | rs17617337 | 1.21E+08 | BAG3 | C | T | 1.06 (1.04–1.08) | 3.65E-09 | C | T | FALSE |
| 12 | rs4766578 | 1.12E+08 | ATXN2 | T | A | 1.04 (1.03–1.06) | 4.90E-08 | T | A | FALSE |
| 16 | rs56094641 | 53806453 | FTO | G | A | 1.05 (1.03–1.06) | 1.21E-08 | A | G | TRUE |

12 independent variants associated with HF at the genome-wide significance level (*P* < 5 × 10−8) (Shah et al 2020). A1 is the reference allele for the expected allelic dosage in UKB. Table indicates which HF SNPs will require dosages to be flipped prior to analysis.

**Web Table 5. Polygenic risk scores examined from the Polygenic Score Catalog.**

| Phenotype | PGS score ID | Number of SNPs in score | Number of score SNPs found in our sample | Percentage of all Score SNPs included in study |
| --- | --- | --- | --- | --- |
| Atrial arrhythmias | PGS000016 | 6,730,541 | 276,790 | 4.1 |
| Diabetes | PGS000014 | 6,917,436 | 282,965 | 4.1 |
| Heavy alcohol intake | PGS000201 | 1,094,954 | 140,902 | 12.9 |
| Hypertension | PGS000706 | 186,726 | 172,158 | 92.2 |
| Myocardial infarction | PGS000710 | 183,566 | 169,242 | 92.2 |
| Obesity | PGS001228 | 27,126 | 24,904 | 91.8 |
| Severe anaemia | PGS001305 | 121 | 98 | 81 |
| Smoking | PGS001129 | 974 | 832 | 85.4 |
| Stable angina | PGS000703 | 183,692 | 169,372 | 92.2 |
| Thyroid disorders | PGS001043 | 69 | 55 | 79.7 |
| Unstable angina | PGS001048 | 687 | 548 | 79.8 |

* PGS score ID is a unique identifier which can be used to lookup details of the score and how it was developed from the PGS catalog (pgscatalog.org). Number of SNPs in score, indicates how many SNPs was included in the PGS score. The last two columns show number and percentage of the score SNPs could be found in our sample, and thus used to calculate the scores with.

**Web Table 6.** Pairwise comparisons (p-values) of survival probability of discovered subtypes in CPRD and THIN data using Log-rank test.

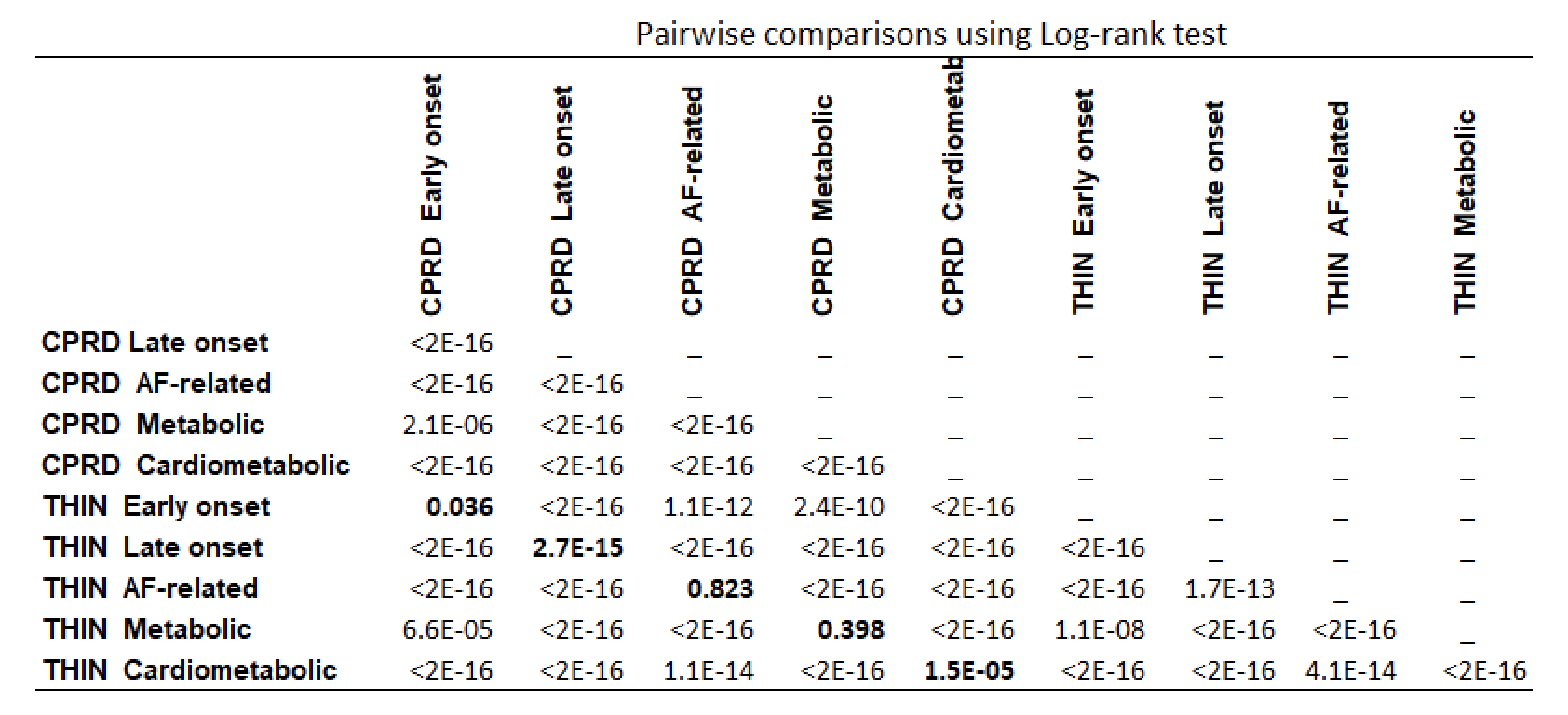
